# Results of a survey of the use of perfusion parameters as a selection tool for pumped deceased donor kidneys in the Organ Procurement Organizations of North America

**DOI:** 10.1101/2024.11.15.24317245

**Authors:** Veerle Heedfeld, Ina Jochmans

## Abstract

**Background:** Hypothermic machine perfusion (HMP) has become a standard method for preserving deceased donor kidneys, offering advantages over static cold storage. Perfusion parameters like renal vascular resistance (RR) have been explored as potential decision-making tools for kidney transplantability, but their clinical use remains unclear.

**Aim:** We aimed to investigate the use of perfusion parameters in decision-making regarding the acceptance of pumped deceased donor kidneys among Organ Procurement Organizations (OPOs) in the USA and Canada.

**Methods:** An anonymous, internet-based survey was sent to 69 OPOs in the USA and Canada, collecting data on the use of HMP, perfusion parameters, and thresholds for transplantability decisions. Descriptive statistics were used for analysis.

**Results:** Of the 67 OPOs contacted, 15 (22%) responded, with 13 complete responses (87%). All OPOs used HMP, with 93% perfusing both donation after brain death and circulatory death kidneys. While 97% of OPOs used perfusion parameters in decision-making, none relied solely on these parameters. Two OPOs (15%) did not use them at all, while six OPOs (46%) considered them with other data or on a case-by-case basis. Only one OPO (9%) reported using specific thresholds for perfusion parameters, applying flow ≥100 mL/min, resistance <0.3 mmHg/mL/min, and pressure between 15–35 mmHg.

**Conclusion:** HMP is widely used, but substantial variability exists in the use of perfusion parameters for transplant decisions. Most OPOs do not rely on these parameters alone and lack standardized thresholds, though specific thresholds are still used.

## Introduction

Hypothermic machine perfusion preservation (HMP), which continuously perfuses a cold acellular preservation solution through the kidneys, has demonstrated advantages over static cold storage (SCS) for all types of deceased donors (donation after brain death (DBD) and DCD).^1^ These benefits include a reduced risk of delayed graft function, cost-effectiveness, and probably better graft survival.^1^

In addition to better outcomes, HMP allows the study of perfusate flows and/or renal vascular resistance (RR) in the kidney. A large body of evidence has investigated whether RR can predict post-transplant outcomes before the kidney is transplanted and we are currently reviewing this evidence in a systematic way.^2^ As such, HMP could be a decision tool assisting the transplant team in the decision whether or not to accept an offered donor kidney for a particular recipient. This evidence shows that there is indeed an independent association of RR on the pump with post-transplant outcomes such as delayed graft function.^3,4^ Nevertheless, the predictive value of RR for a specific kidney-recipient pair is low and not much better than chance.^3,4^

The use of HMP for kidney preservation has gained acceptance over the past two decades with many countries implementing the technology on a standard of individualized basis. In the United States of America, 2023 data form the Scientific Registry of Transplant Recipients (SRTR) shows that 56% of all adult deceased donor kidneys recovered were pumped. The percentage of HMP is higher in kidneys donated after circulatory death (DCD, 76%) than in kidneys donated after brain death (DBD, 44%).^5^ Data from SRTR / Organ Procurement and Transplantation Network (OPTN) also suggest that the use of HMP has decreased deceased donor kidney discard rates.^6,7^ Nevertheless, in 2023 33% of pumped kidneys were not used for transplant after all (34% of DCDs, 31% of DBDs).^5^ It is unclear whether perfusion parameters like RR play a role in the discard of pumped kidneys as SRTR does not collect this specific information.

Indeed, although a large body of evidence has investigated the relationship between RR and post-transplant outcomes, there is little insight in whether or not using RR (or other perfusion parameters) as a decision-making tool is part of clinical practice. We therefore aimed to collect information from Organ Procurement Organizations (OPO) in North America (United States of America and Canada) on the use of perfusion parameters as a decision-making tool in acceptance of pumped deceased donor kidneys.

## Methods

An internet-based, anonymous questionnaire was sent to all OPOs in the USA and Canada (n=69), contacted using the OPOs general email address. The purpose of the questionnaire was briefly explained and participants were asked to answer 6 questions regarding perfusion parameters as a decision-making tool in acceptance of pumped deceased donor kidneys. The full survey is available online.^8^ The survey collected data concerning type of kidneys perfused, incentives to pump, perfusion parameters as a tool for acceptance of kidneys and the threshold to consider kidneys for transplant. Study data were collected and managed using REDCap electronic data capture tools hosted at KU Leuven.^9,10^ Data are presented with descriptive and summary statistics.

This study has been presented to the Privacy Committee of KU Leuven who deemed that a formal advice was not needed because this is not a clinical study nor are personal data being collected. The protocol and survey questions were reviewed and approved by the Ethical Committee – Research of UZ Leuven / KU Leuven (S69499).

## Results

### Survey participants

The survey was distributed on September 19, 2024 to 69 OPOs operating in the USA and Canada. It remained open for three weeks, with two reminders sent to non-responders on September 23 and 30, 2024. Eleven email delivery failures were identified (11/69; 16%) and alternative contact methods were used in nine cases: seven OPOs were reached via online webforms, and two through alternative email addresses. For the remaining two cases, no electronic contact details could be located. Thus, the survey successfully reached 67 OPOs (67/69; 96%).

Among these, 15 responded (15/67; 22%), yielding 13 complete responses were received (13/15; 87%), one partial response (1/15; 6.5%) and one response with no submitted data (6.5%). The partial response included only an answer to whether kidneys were pumped but did not provide additional data. This case was excluded from the analyses of subsequent responses. The response with no data was also excluded from further analyses.

### Reasons to pump kidneys

All responding OPOs reported that kidneys were pumped (14/14; 100%). In the majority of cases, all deceased donor kidney types (DBD and DCD) were pumped (13/14; 93%), while one OPO reported pumping only DCD kidneys (1/14; 7%).

Thirteen OPOs provided reasons for kidney pumping (13/14; 97%), which included: adherence to standard policy (4/13; 31%), anticipated long cold ischemia time (8/13; 62%), kidneys with a Kidney Donor Profile Index (KDPI) >85% (5/13; 38%), and other reasons (8/13; 62%) (Fig. 1, Table 1). All OPOs used the LifePort® device from Organ Recovery Systems (Itasca, IL, USA) for kidney pumping (13/13; 100%).

**Table 1.** Other reasons listed to pump deceased donor kidneys.

| Other reasons for pumping kidneys | OPOs that listed this reason; n (%) |
| --- | --- |
| High creatinine or acute kidney injury | 5 (38%) |
| Poor organ flush | 2 (15%) |
| Long warm ischemia time | 1 (8%) |
| Combined organ transplant (e.g. liver and kidney) | 1 (8%) |
| At the request of the transplanting surgeon | 2 (15%) |
| “Marginal kidneys” for a combination of reasons | 1 (8%) |
| Donor age | 2 (15%) |

*Multiple reasons are possible so the sum does not add up to 100%*

**Fig. 1.**
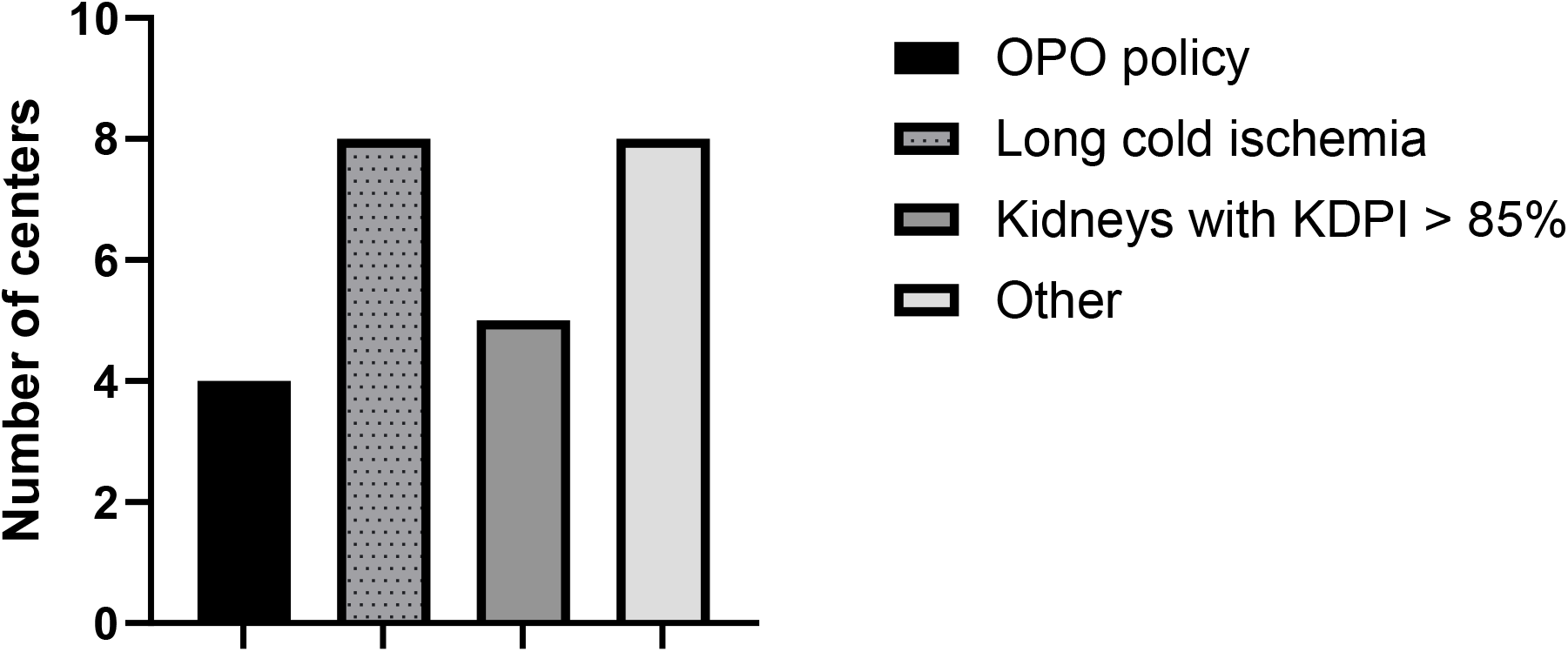
Reasons to pump kidneys KDPI, kidney donor profile index; OPO, organ procurement organization

### Use of perfusion parameters as decision tool

Of the OPOs that provided information on the use of perfusion parameters (13/14; 97%), none reported using these parameters as standalone criteria for decision-making. Two centers indicated they do not use perfusion parameters in their decision-making process at all (2/13; 15%). In six centers, perfusion parameters are either considered in conjunction with other clinical data (6/13; 46%) or used on a case-by-case basis (6/13; 46%) (Fig. 2).

**Fig. 2.**
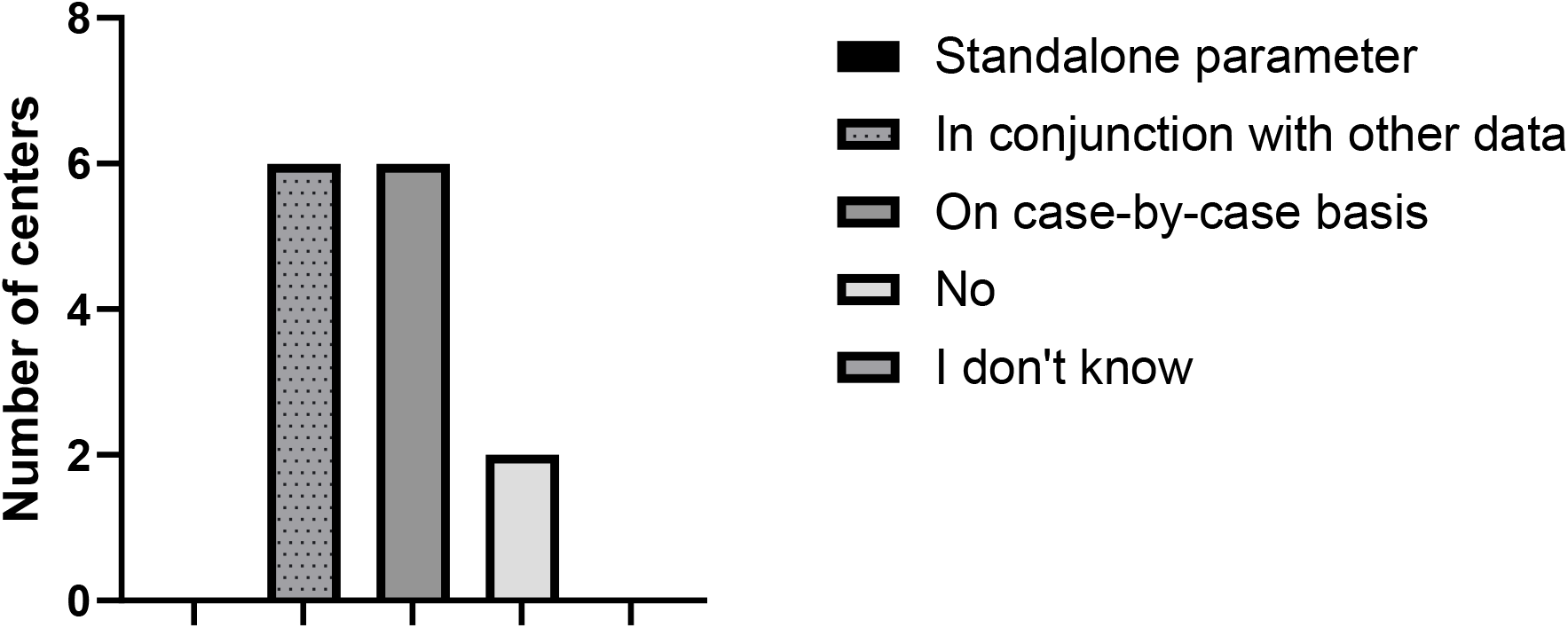
Use of perfusion parameters in decision whether kidney is transplantable

When asked about the use of thresholds to determine kidney transplantability to the 11 centers who use perfusion parameters in their decision-making, two centers reported being unsure (2/11; 18%). Eight OPOs stated they do not use fixed thresholds but instead assess transplantability on a case-by-case basis (8/11; 73%) (Fig. 3). Only one center reported applying specific thresholds for perfusion parameters to determine transplantability (1/11; 9%). The thresholds used by this center are: a flow of ≥100 mL/min, RR of <0.3 mmHg/mL/min, and pressure between 15–35 mmHg.

**Fig. 3.**
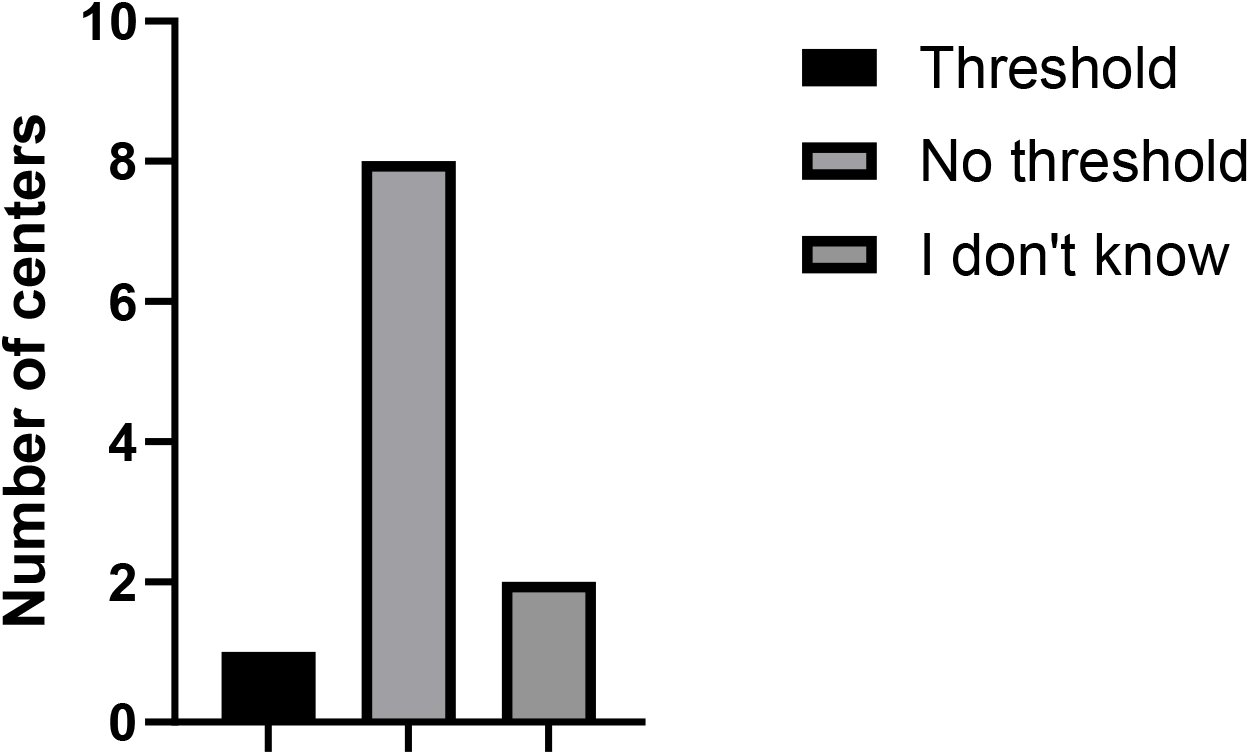
If perfusion parameters are used as a decision-making too, is a threshold value of these perfusion parameters used?

## Discussion

This study provides valuable insights into the current practices of OPOs in the USA and Canada regarding the use of HMP and perfusion parameters in the assessment of deceased donor kidneys. While HMP has become an established method for organ preservation, the extent to which perfusion parameters influence decision-making for kidney transplantation remains unclear. Our findings highlight significant variability and uncertainty in the application of these parameters as decision-making tools, suggesting a need for further research and potential standardization.

The data show that perfusion parameters are not widely used as standalone criteria for determining kidney transplantability. This aligns with the evidence suggesting that while RR is associated with post-transplant outcomes such as DGF, its predictive value remains limited when applied in isolation. Instead, many OPOs incorporate perfusion parameters into a broader context that includes clinical and donor-specific factors. The use of perfusion parameters on a case-by-case basis (46%) or alongside other data (46%) underscores the complexity of decision-making in organ transplantation and the lack of consensus on the role of these metrics. However, there is still evidence that thresholds are used in decision-making. The thresholds used are consistent with values cited in the literature, but their isolated application is unlikely to capture the full complexity of organ viability. Most OPOs either lacked fixed thresholds (73%) or were unsure of their usage (18%).

The reliance on case-by-case decision-making suggests that transplant teams prefer individualized assessments that incorporate a combination of clinical judgment, donor characteristics, and perfusion data. However, this approach may introduce variability and subjectivity into decision-making, potentially impacting organ utilization rates.

There are limitations to this study. The response rate for this survey limits the generalizability of our findings. While the responding OPOs represent a diverse sample, it is possible that non-responders have different practices regarding the use of perfusion parameters. Additionally, the small sample size for certain questions, such as the use of thresholds, restricts the robustness of our conclusions.

In conclusion, this study highlights the widespread use of HMP among OPOs and the variability in how perfusion parameters are employed in decision-making for kidney transplantation. While these parameters are often considered alongside other clinical data, their use as standalone criteria or with defined thresholds remains still happens.

## Data Availability

All data produced in the present work are contained in the manuscript.

https://osf.io/6yvp7/

## Acknowledgements

We would like to thank all survey participants. We acknowledge the use of ChatGPT, developed by OpenAI, for assisting with the drafting and refinement of this manuscript. As experienced researchers, we were curious to explore how AI could contribute to the writing process, and the support proved valuable in enhancing the clarity of the paper.

## Declarations

IJ received an unrestricted research grant from XVIVO Perfusion paid to her institution.

## Data availability statement

All data produced in the present work are contained in the manuscript.

## Funding statement

This study did not receive any funding.

## Notes

### Clinical Protocols

https://osf.io/6yvp7/

### Author Declarations

Ethics Committee REsearch of UZ/KU Leuven gave ethical approval for this work (reference number S69499).

